# Practices of midwives in the management of postpartum haemorrhage: A case of Maseru, Lesotho

**DOI:** 10.1101/2024.02.16.24302896

**Authors:** Mpho Chabeli, Isabel Nyangu, Regina Mpemi

## Abstract

Postpartum haemorrhage (PPH) remains the common cause of maternal mortality. More than half of maternal deaths from PPH occur within 24 hours of delivery and could be prevented through midwives’ compliance with guidelines and standards for clinical practice. This study aimed to assess the midwives’ practices regarding the management of PPH in Maseru Lesotho. A quantitative cross-sectional study was undertaken. Convenience sampling was used to select 220 midwives who voluntarily completed a structured self-administered questionnaire. Ethical clearance was granted by the Ministry of Health Research and Ethics Committee. Data was analysed using Statistical Package for Social Science and presented using descriptive and analytic statistics. Midwives were competent with estimation and recording blood loss, placenta assessment and vital signs of patients. There was a significant association between the highest education qualification and midwives who estimate and record blood loss, palpate and measure the uterine fundus and assess the completeness of the placenta. The results revealed a significant association between work experience, uterine fundus measurement and estimated blood loss. Midwives reported to practice management of PPH based mostly on guidelines. However, many midwives still disregard recommendations, putting patients at risk hence the need to intensify supervision to ensure safe practices.

## INTRODUCTION

Midwives play an important role in preventing, early detection and management of postpartum haemorrhage (PPH) through careful observation of the mother (Mohamed et al., 2019.). Postpartum is a critical time with the potential for complications as bleeding can rapidly become life-threatening (Romano et al., 2010). Yevoo et al. (2020) further emphasise that midwives should adhere to guidelines for clinical practice to ensure better health outcomes for the mother and baby. Should the recognition of PPH be late, it will pose a great hindrance to management, even in highly resourced settings (Chau & Farber, 2020).

Globally, 303,000 women are estimated to be dying from avoidable obstetric causes related to pregnancy and childbirth each year and 66% of those deaths were from Sub-Saharan Africa (Okonofua et al., 2019). According to Khan et al. (2019), 14 million women around the world suffer from PPH every year and it accounts for approximately 35% of all maternal deaths in both developed and developing countries (Aboelhadedl et al., 2019.).

In Lesotho, the maternal mortality rate in 2014 was 1024 per 100,000 live births and more than 40% of maternal mortality occurred in the district of Maseru out of 10 districts (Ministry of Health, Lesotho, 2016). Of all these deaths, the dominant cause was PPH at a proportion of 31.4% (Mashea et al., 2018.).

For successful implementation of maternal healthcare guidelines, midwives should be capacitated through training, supported, and supervised to execute PPH management with ease (Ramavhoya et al., 2021). Panyapin (2020) established that the majority of midwives had implemented the recommendations of guidelines for the management of PPH in their practice, however, Ahmed Elkholy et al. (2017) discovered that most studied midwives did not perform some procedures such as urinary catheterization and wound care, and less than three-quarters of nurses performed practices such as (fundus assessment, lochia assessment, episiotomy care, pulse, blood pressure and IV fluid). These findings clarify the lack of regular training courses and sessions by the Ministry of Health and other health authorities, they highlight the need to improve the content of the nursing curriculum together with continued training courses which yield well-trained and educated nurses about the management of PPH. The findings of this study disagree with Faiza(2015) who reported that the practical aspects of midwives regarding the management of PPH were (69.6 %), such as (assessment of fundal level 73.3%, importance of empty bladder 75%, assessment of lochia and blood loss 61.7%, vital signs 65.5% and IV fluid 93.9 %).

Angelina et al. (2019) found that midwives demonstrated inadequate skills for the management of PPH. This is not in agreement with findings from Ogundeko et al. (2017.) who found that about 60% of participants managed PPH at various primary care centres in Nigeria. Among the management skills for prevention and control of PPH stated, they included early cord clamping, emptying of bladder during the third stage of labour and use of uterotonics such as oxytocin. This agrees with UNICEF and WHO (2019.) outline guidelines for the management of PPH whenever a midwife is assisting with the delivery. Those include active management in the administration of uterotonics soon after the birth of the baby, clamping of the cord following the observation of the uterine contraction and delivery of the placenta by controlled cord traction, following uterine massage.

It is assumed that evidence-based guidelines for the management of PPH are not optimally adhered to, leading to substandard care and a possible cause for inadequate reduction of morbidity and death owing to PPH (de Visser et al., 2018). Midwives, therefore, play an important role in the prevention and early recognition of PPH, especially during the early postpartum period through careful observation of the mother (Mohamed et al., 2019.).

This study was prompted by the fact that despite the existence of guidelines developed by the Government of Lesotho through the Ministry of Health for the management of PPH, it continues to be the top cause of maternal deaths.

## RESEARCH METHOD

A quantitative cross-sectional study was undertaken to assess the practices of midwives regarding the management of PPH in Maseru, Lesotho. A total of 220 midwives who were conveniently sampled from various healthcare facilities participated in the study by completing a structured self-administered questionnaire. Written informed consent was sought from the participants who voluntarily took part. They were allowed to ask questions and could withdraw from the study without any prejudice. Permission to conduct the study was sought and granted by the National University of Lesotho Institutional Review Board and the Ministry of Health Research and Ethics Committee (ID-198-2022). Gatekeeper permission was also sought from the facility managers.

## RESULTS AND DISCUSSIONS

### Demographic characteristics

There was a total of 220 participants in this study of which the majority were females 80% (n=178) and the dominating position held by the participants was a Nursing sister (a title for entry-level midwives) 78% (n=172). The educational level attained by most participants was Diploma 75% (n=166) followed by Degree 19% (n=41). Most of the participants 69% (n= 151) reported to have not attended an in-service training on guidelines on PPH. Participants with over 7 years 45% (n=99) of experience dominated followed by those with 1-3 years 30% (n=66). More than half of the participants were working in health centres 55% (n=122).

### Practices of midwives for management of PPH during the fourth stage of labour

Estimation and recording of blood loss in the immediate post-partum phase is an essential practice for nurse-midwives as it enables them to assess the status of a woman who has just delivered. There were 124 (56.4%) midwives who rated themselves as excellent with estimation and recording of blood loss and only 6 (2.7%) rated themselves as average. About the assessment of the completeness of the placenta, 69.5% (n=153) rated themselves to be excellent,) 25.5% (n=56) rated themselves very good and 5% (n=11) were rated Good. On checking the vital signs of the patients, 182 (82.7%) of midwives rated themselves as excellent and 0.5% (n=1) rated him/herself as average.

Presented in Table 1.2 is the Chi-square distribution of midwives’ practices towards the management of PPH and demographic characteristics. The results suggest a significant association between the highest education qualification and the midwives who estimate and record blood loss [X^2^(6, N= 220) = 26.2, p<0.05], midwives who palpate and measure the uterine fundus [X^2^(6, N= 220) = 21.905, p<0.05]. There was also a significant association between the Highest qualification and nurses who assess the completeness of the placenta [X^2^ (4, N= 220) = 23.97, p<0.05].

**TABLE 1.1:** Nurse midwives’ ability to carry obstetric care Practices for the Prevention of PPH during the fourth stage of labour

| Statement: | FREQUENCY AND PERCENTAGES OF PRACTICES SELF-RATING |  |  |  |  |  |  |  |  |  |
| --- | --- | --- | --- | --- | --- | --- | --- | --- | --- | --- |
|  | Excellent |  | Very good |  | Good |  | Average |  | Below average |  |
| Ability to estimate and record blood loss | 124 | 56.4% | 65 | 29.5% | 25 | 11.4% | 6 | 2.7% | 0 | 0.0% |
| Ability to palpate and measure the uterine fundus | 160 | 72.7% | 50 | 22.7% | 8 | 3.6% | 2 | 0.9% | 0 | 0.0% |
| Ability to assess completeness of placenta | 153 | 69.5% | 56 | 25.5% | 11 | 5.0% | 0 | 0.0% | 0 | 0.0% |
| Ability to examine the perineum for cervical or vaginal tears, or episiotomy | 176 |  | 37 | 16.8% | 6 | 2.7% | 1 | 0.5% | 0 | 0.0% |
| Ability to estimate blood loss through examination of lochia amount and consistency | 128 | 58.2% | 62 | 28.2% | 19 | 8.6% | 10 | 4.5% | 1 | 0.5% |
| Ability to check vital signs | 182 | 82.7% | 32 | 14.5% | 5 | 2.3% | 1 | 0.5% | 0 | 0.0% |
| ability to complete the records/recording | 156 | 70.9% | 44 | 20.0% | 20 | 9.1% | 0 |  | 0 | 0.0% |
| Ability to encourage the woman to empty the bladder | 164 | 74.5% | 45 | 20.5% | 11 | 5.0% | 0 | 0.0% | 0 | 0.0% |
| Ability to provide health education on danger signs of the postpartum period | 158 | 71.8% | 42 | 19.1% | 17 | 7.7% | 2 | 0.9% | 1 | 0.5% |
| Ability to keep the woman for 12 hours in a facility | 134 | 60.9% | 47 | 21.4% | 23 | 10.5% | 15 | 6.8% | 1 | 0.5% |

**TABLE 1.2:** Chi-Square distribution of midwives’ practices towards management of PPH, Highest Qualification and work experience

| Statement:<br>I would: | Highest Qualification |  |  | Work Experience |  |  |
| --- | --- | --- | --- | --- | --- | --- |
|  | Pearson Chi-Square | df | Asymptotic Significance (2-sided) | Pearson Chi-Square | df | Asymptotic Significance (2-sided) |
| Estimate and record blood loss | 26.242<sup>a</sup> | 6 | 0.000 | 10.280<sup>a</sup> | 6 | 0.113 |
| Palpate and measure the uterine fundus | 21.905<sup>a</sup> | 6 | 0.001 | 13.069<sup>a</sup> | 6 | 0.042 |
| Examine the completeness of the placenta | 23.967<sup>a</sup> | 4 | 0.000 | 9.186<sup>a</sup> | 4 | 0.057 |
| Examine the perineum for cervical or vaginal tears, or episiotomy | 2.430<sup>a</sup> | 6 | 0.876 | 3.169<sup>a</sup> | 6 | 0.787 |
| Estimate blood loss through examination of lochia amount and consistency | 11.637<sup>a</sup> | 8 | 0.168 | 16.009<sup>a</sup> | 8 | 0.042 |
| Check vital signs | 7.112<sup>a</sup> | 6 | 0.311 | 7.348<sup>a</sup> | 6 | 0.290 |
| Complete the records/recording | 15.491<sup>a</sup> | 4 | 0.004 | 5.043<sup>a</sup> | 4 | 0.283 |
| Encourage the woman to empty the bladder | 10.864<sup>a</sup> | 4 | 0.028 | 11.228<sup>a</sup> | 4 | 0.024 |
| Provide health education on danger signs of the postpartum period | 7.886<sup>a</sup> | 8 | 0.445 | 5.322<sup>a</sup> | 8 | 0.723 |
| Keep the woman for 12 hours in a facility | 10.332<sup>a</sup> | 8 | 0.242 | 12.007<sup>a</sup> | 8 | 0.151 |

The results further revealed that there was a significant association between the work experience of the midwives and how they palpate and measure the uterine fundus [X^2^ (6, N= 220) = 21.91, p<0.05], also how they estimate blood loss through examination of lochia amount and consistency [X ^2^ (8, N= 220) = 11.637, p<0.05].

The findings of the study have revealed that most of the participants were within the age group of 31-35(27%), thus showing that the population was young, similar to the study conducted by Tsiame (2018.) which showed that the majority of participants were younger than 40 years. The findings of this study indicated that the majority of participants, 81% were females. These results are consistent with some published studies that have shown that females tend to dominate the nursing profession (Muzeya, 2015; Sethi et al., 2019).

Accurate assessment and recording of blood loss, monitoring of maternal vital signs, fluid replacement, and arrest of any of the source of haemorrhage as recommended by PPH guidelines, in the immediate post-partum are essential practices for midwives as this enables them to assess the status of a woman who has just delivered (Bienstock et al., 2021). The findings of the present study reveal that more than 50% of midwives (56.4%) reported estimating and recording blood loss during postpartum as recommended by the guidelines. The results suggest a significant association between the Highest Education Qualification and the midwives who estimate and record blood loss (p=0.000). However, this finding disagrees with the finding of Sattarzadeh et al. (2017) who found no significant association between the highest qualification and the practices of midwives.

Regarding the assessment of completeness of the placenta, 69.5% of midwives reported assessing completeness of the placenta and membranes during their practice. Similarly, Wake & Wogie (2020) in their study conducted in government health institutions in Tigray Region Northern Ethiopia indicated that most midwives (67.3%) reported assessing the completeness of the placenta and membranes during their practice. Likewise, Faiza (2015) found that 76.7% of midwives reported checking parts and membranes after placenta delivery. However, the results of the present study are much higher than those of (Mohammad, 2019.), who found that 26.1% of midwives reported examination of the placenta and membranes for completeness as part of practice in the management of PPH.

Concerning checking the vital signs of the patients, 82.7% of midwives reported checking patients’ vital signs as per guidelines. These findings agree with Chimtembo et al. (2013), in their study conducted in the Dedza district in Malawi, who found that 62% of the midwives reported that they monitored mothers’ conditions at least once during the fourth stage of labour. The findings of the current study further agree with (Mohammad, 2019.) who established that 74.1% of midwives reported checking vital signs of patients during the postpartum period.

This study found that the majority of midwives (80%) reported examining the perineum for cervical or vaginal tears, or episiotomy in line with the guidelines. Likewise, Wake & Wogie (2020) in their study conducted in government health institutions in Tigray Region Northern Ethiopia, indicated that most midwives (74.5 %) reported assessing maternal genitalia for tear and trauma. The findings of the current study were much higher than the result of the study conducted by Chimtembo et al. (2013) in the Dedza district in Malawi, who found that 48% of midwives reported conducting perineal inspection. (Faiza (2015), also found results that are lower than those of the current study, which found that 63.3% of midwives reported inspection of the perineum after delivery as part of their practice during postpartum. Similarly, Mohammad (2019.) found that all midwives included in his study reported examining the utero-vaginal canal and perineum for trauma and prompt repair if present. This difference might be brought about by a lack of supervision and mentorship especially from the Ministry of Health to ensure adherence of midwives to guidelines.

The present study has shown that 58.2% of midwives had reported estimating blood loss through examination of lochia amount and consistency. This finding is similar to Faiza (2015), who established that 61.7% of midwives in his study reported assessment of blood loss and assessment of lochia as part of their practice for the management of PPH. The results of the current study are lower than those of Muhamed (2019) who has shown that 77.8% of midwives in his study included examination of lochia for amount and consistency in their practice for management of PPH. This difference might be a result of a lack of in-service training and poor supervision and mentorship programs and because of newly deployed midwives who had not been oriented or supervised on the management of PPH. The results further revealed that there was a significant association between the work experience of the midwives and midwives who estimate blood loss through examination of lochia amount and consistency (p=0.042). However, this finding is not consistent with the finding of Nguyen et al. (2020) who established no correlation between work experience and practice.

The current study revealed that a significant number of midwives (70.9%) reported completing patients’ records. This is not in line with the results of Mutshatshi et al.(2018) who conducted a study in South Africa and revealed that midwives did not complete patient records and some information was never recorded. This study further differs from Juwita et al. (2019) in their study conducted in Indonesia which established that a significant number of midwives did not complete the data on the patients’ management during postpartum owing to work overload and lack of time to complete the records.

The present study established that the majority of midwives (74.5%), reported encouraging the woman to empty the bladder. This finding is in agreement with Faiza (2015) in his study conducted at three teaching hospitals in -Khartoum State-Sudan, who revealed that 75% of midwives encouraged the woman to empty the bladder. The current study results differ from (Mohammad, 2019.) who discovered that only 40.7% of midwives reported encouraging the mother to evacuate her urinary bladder regularly. This difference could be an impact of staff rotations where midwives have no experience and have not been trained in the management of PPH.

About providing health education on danger signs of the postpartum period 71.8% of midwives reported providing health education as part of their practice during the postpartum period. Similarly, Chimtembo et al.(2013), in their study conducted in the Dedza district in Malawi, found that many midwives (69%) reported having imparted information on danger signs during the postpartum period. However, this finding differs from Faiza (2015) who found much lower results (30%) of midwives reported to have provided education to women on danger signs during postpartum.

It is encouraged that the woman and the newborn should not be discharged earlier than 24 hours postpartum because a bulk of maternal deaths occur within the first twenty-four hours after childbirth (WHO, 2022). The current study however revealed that 60.9% of midwives reported to keep the woman for 12 hours at the facility. This could be related to a lack of infrastructure unavailability of food supplies and understaffing. Reducing the time for the provision of postpartum care significantly contributes to poor maternal outcomes as interventions for preventing morbidity and mortality related to PPH will not be timely. The Ministry of Health is therefore encouraged to extend maternity wards and ensure that they are fully staffed.

Accurate assessment, monitoring of the maternal condition and recording of blood loss, post-delivery are recommended by PPH guidelines so that any signs of haemorrhage and arrest of the sources, thereof, are implemented timeously before complications could arise. The midwives therefore implemented these as per guidelines that they should check the vital signs of patients (82.7%); encourage the woman to empty the bladder (80%); provide health education on danger signs of the postpartum period (74.5%); complete patients’ records (71.8%); assess the completeness of placenta and membranes (70.9%) and estimate and record blood loss during postpartum (69.5%).

However, it has been noted that a percentage range of about 18% to 44%, translating to about 40 to 96 participants, were not completely adherent to the stipulation of the PPH guidelines. This is of serious concern as it indicates that many women are still at risk of substandard care thus leading to their untimely demise attributed to preventable courses. The second delay, delay in initiating treatment, can be regarded as the main cause of maternal mortality, even when women manage to seek help on time. This could be the reason for the continued high levels of morbidity and mortality despite high ANC (77%) attendance and increasing facility-based delivery from 68% in 2018 to 74% in 2019 (MOH, District Health Information System2). These, therefore, corroborate the significance of quality intrapartum care and continued skilling of midwives through the provision of in-service training. It is worth noting that a significant majority (69%) of participants mentioned that they did not receive training on the use of PPH guidelines.

## CONCLUSION

The results of the study demonstrated that midwives reported to practice management of PPH based on guidelines to a very high extent. However, there is still a significant number of midwives who do not comply with the precepts of the PPH guidelines consequently putting patients at risk hence the need to intensify supervision to ensure safe practices. This, therefore, requires urgent attention, if the country is to reduce the current trends of maternal mortality to less than 70 per 100,000 live births by 2030 as per SDG (3.1). This is mainly because even one death of a mother is unacceptable.

## Data Availability

All data produced in the present study are contained in the manuscript.

## ACKNOWLEDGEMENTS

The researcher acknowledges the supervisors and midwives who participated in the study.

